# Virtual reality-assessment of social interactions and prognosis in depression

**DOI:** 10.1101/2022.09.28.22280445

**Authors:** Suqian Duan, Lucia Valmaggia, Andrew J. Lawrence, Diede Fennema, Jorge Moll, Roland Zahn

## Abstract

**Importance:** Stratification of depression for personalised treatment is urgently needed to improve poor outcomes. Excessive self-blame-related motivations such as self-punishing tendencies have been proposed to play a key role in the onset and maintenance of depression. Their prognostic role, however, remains elusive.

**Objective:** Use Virtual Reality (VR) to determine whether maladaptive self-blame-related action tendencies are associated with a poor prognosis for depression when treated as usual in primary care (pre-registered: <u>NCT04593537</u>).

**Design:** Remote prospective cohort study (6/2020-6/2021) with four months follow-up.

**Settings:** Online recruitment from primary care and self-report.

**Participants:** n=879 pre-screened, n=164 eligible, n=101 completed baseline (age:18-66 years, mean=32.05±12.32, n=89 female), n=98 the VR-task, and n=93 the follow-up. Main inclusion criteria: at least one antidepressant medication trial and Patient Health Questionnaire-9≥15 at screening; main exclusion criteria: screening above threshold on validated self-report instruments for bipolar or alcohol/substance use disorders.

**Exposure(s):** All participants completed a VR assessment via headsets sent to their homes, as well as online questionnaires to measure their clinical characteristics.

**Main outcomes and Measures:** Primary: Quick Inventory of Depressive Symptomatology self-reported-16 score after four months. Hypotheses in the study were formulated before the data collection and pre-registered.

**Results:** Contrary to our specific prediction, neither feeling like hiding nor creating a distance from oneself was associated with prognosis of depression during the follow-up period in the pre-registered regression model. Using a data-driven principal components analysis of all pre-registered continuous measures, a factor most strongly loading on punishing oneself for other people’s wrongdoings (β=.23, p=.01), a baseline symptom factor (β=.30, p=.006) and Maudsley Staging Method treatment-resistance scores (β=.28, p=.009) at baseline predicted higher depressive symptoms after four months. This relationship was confirmed by a significant interaction between feeling like punishing oneself for others’ wrongdoings and time of monthly follow-up which was driven by higher depressive symptoms at last follow-up [F(1,84)=6.45, p=.01, partial Eta Squared=0.07] in the subgroup who had reported feeling like punishing themselves at baseline. Our pre-registered statistical learning model prospectively predicted a cross-validated 19% of variance in depressive symptoms.

**Conclusions and Relevance:** Feeling like punishing oneself is a relevant prognostic factor and should therefore be assessed and tackled in personalised care pathways for difficult-to-treat depression.

**Key points:** *Question:* Can remote virtual-reality (VR)-based measures of social interactions be used to identify risk factors of poor prognosis in depression?

*Findings:* In this pre-registered remote prospective cohort study in 101 participants with depression and follow-up over four months, the VR-assessed feeling like punishing oneself for other people’s wrongdoing was the sole prognostic risk factor apart from known clinical variables such as symptom severity and previous treatment-resistance.

*Meaning:* Feeling like punishing oneself is a relevant prognostic factor and should therefore be assessed and targeted in difficult-to-treat depression.

## Introduction

Only a third of patients reach remission after their initial treatment (e.g. Rush et al., 2008) and multiple treatment-gaps at each stage of the care pathway for depression have been identified (Strawbridge et al., 2022). The ability to stratify depression by likely prognostic trajectory early on, could accelerate access to more personalised treatments in a cost-effective way. Yet, scalable measures with prognostic relevance are scarce and insufficient for stratification. Freud described self-blaming feelings and an implicit need for self-punishment as distinctive features of depression (Freud, 1917). The latter was criticised by Beck as limiting the psychoanalytical approach to depression by implying people with depression were not motivated to get better (Beck, 1985). This may be one of the reasons why the focus of empirical research over the past decades has been on self-blaming emotions in depression (O’Connor et al., 2002; Zahn et al., 2015) rather than systematically characterising the associated implicit action tendencies (e.g. Janoff-Bulman, 1979; Tangney et al., 2007), such as feeling like punishing oneself or hiding. Action tendencies which precede social actions, are likely to play an evolutionary important role in the social survival of human beings (e.g. Darwin, 1872; Haidt, 2003). Their role in the prognosis of current depression, however, is elusive.

Action tendencies are essential components in appraisal theories of emotions (Frijda et al., 1989; Moors, 2009; Roseman et al., 1994). Self-blame-related action tendencies (e.g. feeling like hiding) are the motivational component of self-blaming emotions (e.g. shame), which could play either adaptive or maladaptive roles (Tangney et al., 2007). Gray’s reinforcement sensitivity theory (Gray, 1970), proposes two dissociable neural systems as dimensions of human action tendencies: behavioural “activation” and “inhibition” – the latter is also referred to as “withdrawal”. Stronger behavioural inhibition/withdrawal and impaired behavioural activation were associated with affective disorders (e.g. Kasch et al., 2002). In line with this, Tangney et al. (2007) postulated that adaptive self-blame-related action tendencies are more associated with behavioural activation and involve proactive pursuit such as reparative actions, whereas maladaptive self-blame-related action tendencies motivate social withdrawal and interpersonal separation (Duan, Lawrence, et al., 2022; Tangney et al., 2007). In keeping with this hypothesis, using a text-based task, we recently showed that withdrawal-related maladaptive action tendencies including hiding and creating a distance from oneself were more pronounced in patients with remitted depression compared with control participants (Duan, Lawrence, et al., 2022) and were associated with subsequent recurrence risk (Duan, Lawrence, et al., 2022; Duan, Valmaggia, et al., 2022).

For the present prospective study, we used a novel VR task developed and validated in our separately reported cross-sectional study comparing people with and without depression at baseline to examine our pre-registered hypothesis 1 (NCT04593537). The cross-sectional study revealed that people with depression showed a distinctive tendency to attack/punish themselves, which was specifically associated with a history of self-harm but not suicide attempts (Duan, Valmaggia, et al., 2022). Here, we prospectively probed the prognostic role of maladaptive self-blame-related action tendencies for depressive symptoms over four-months of follow-up whilst being treated as usual in primary care. Previous measures of action tendencies used abstract descriptions of social scenarios which limited engagement in the scenarios and ecological validity (Duan, Valmaggia, et al., 2022; Mu & Berenbaum, 2019). Our immersive VR task made it possible to act out the action tendencies as well as to increase the engagement of participants. We hypothesized that maladaptive self-blame-related action tendencies at baseline are associated with a poor prognosis for current major depression when measured four months after baseline (pre-registered hypothesis 2). Specifically, based on our previous findings (Duan, Lawrence, et al., 2022), in our pre-registered analysis plan, we highlighted two withdrawal-related maladaptive action tendencies (hiding and creating a distance from oneself), which we predicted to be associated with poor prognosis of depression. In addition, based on the wider clinical literature, we pre-registered a number of clinically established risk factors, chiefly measures of depressive and anxiety symptoms, as well as treatment-resistance as further detailed in the methods section. Our pre-registered research question 3 was whether maladaptive self-blame-related action tendencies can be used to predict prognosis in MDD at the individual level when combined with other predictors using a nested elastic-net regularised doubly-cross-validated regression model [dCVnet, https://github.com/AndrewLawrence/dCVnet, (Lawrence et al., 2021)).

## Methods

### Participants

This study was approved by the King’s College London PNM Research Ethics Subcommittee (Project Reference:HR-19/20-17589) and pre-registered (NCT04593537) prior to data collection. All participants were recruited via online advertising. Participants were compensated with a £25 Amazon voucher on completing the study or £15 for only completing the online baseline assessment. Participants’ eligibility was assessed by an online pre-screening questionnaire. The inclusion criteria were: age ≥ 18 years; at least moderately severe major depressive syndrome [The Patient Health Questionnaire-9 (Kroenke et al., 2001) score ≥ 15]; early treatment resistance to antidepressants as defined as one antidepressant medication treatment trial in primary care (Fekadu et al., 2018) and being able to complete self-report scales orally or in writing. Exclusion criteria were: a previously diagnosed or likely bipolar disorder [Hypomanic Checklist-16 (Forty et al., 2010) score > 8, with symptoms lasting ≥ 2 days, and endorsing two of the first three screening questions of the Composite International Diagnostic Interview bipolar screening scale (Kessler et al., 2006)], a personal history of schizophreniform symptoms [three clinical screening questions to exclude schizophreniform disorders (Lythe et al., 2015)], drug or alcohol abuse over the last 6 months [Primary Care Evaluation of Mental Disorders (Spitzer et al., 1994), modified to screen for drug abuse], a suspected central neurological condition, a planned or current pregnancy, or currently being treated by a mental health specialist in secondary care. More information about the inclusion/exclusion reasons can be found in the Supplementary Materials of Duan, Valmaggia, et al. (2022).

In total, 879 participants completed the pre-screening questionnaire, of which 164 were eligible. Of those, 101 consented to and took part in the study. All 101 participants completed the baseline assessment and 98 of them completed the VR task. Participants completed follow-up assessments at one (n=94), two (n=91), three (n=80) and four (n=93) months (see Supplementary Table 1 and 2 for demographic and clinical characteristics).

### Assessments of clinical characteristics

All demographic information and clinical characteristics of participants were collected using online self-report.. Participants’ depressive symptoms were measured by the Quick Inventory of Depressive Symptomatology (self-reported, 16 items) (QIDS-SR-16, Rush et al., 2003) as our primary outcome at baseline and four months’ follow-up only and by the Very Quick Inventory of Depressive Symptomatology (VQIDS-SR-5, De La Garza et al., 2017)] at one, two and three months’ follow-ups. Other clinical characteristics collected include anxiety symptoms as measured by the Generalised Anxiety Disorder -7 Scale (GAD-7) (Spitzer et al., 2006), information about participants’ depressive episodes as well as their current and past antidepressant medications. We also used the Maudsley Modified Patient Health Questionnaire-9 (Harrison et al., 2021) as a secondary outcome. More details on assessments of clinical characteristics can be found in Duan, Valmaggia, et al. (2022).

### Virtual reality assessment of blame-related action tendencies

The virtual reality assessment of blame-related action tendencies, deployed on Oculus Go devices, was developed based on the value-related moral sentiment task (Zahn et al., 2015) and has been described and validated in our previous cross-sectional study (Duan, Valmaggia, et al., 2022). There were 30 scenarios in the task, in which either the participant (self-agency condition, 15 scenarios) or the participants’ friend (other-agency condition, 15 scenarios) acted counter to social and moral values in hypothetical social interactions between the participants and their friends. In each scenario, participants were taken to a scene (e.g. in the street or a shopping centre) and saw a VR avatar (their friend) moving towards them, while a narrator described the hypothetical social scenarios. Participants were then asked to choose an action they felt like doing from choice options displayed on the screens: “feeling like verbally attacking my friend”, “punishing myself”, “apologising”, “hiding”, “creating distance from my friend”, “creating distance from myself” and “other/no action”. After participants made their choices, the display changed accordingly to act out the corresponding actions. At the end of the scenarios, participants were asked to rate their levels of self-blame and other-blame from 1 (not at all) to 7 (very much), shown on the screen as visual analogue scroll bars. For details, videos and screenshots of the VR task, see Duan, Valmaggia, et al. (2022).

### Procedure

This study was conducted fully remotely. After participants consented to the study, they received the link to the baseline assessment and email instructions to complete the VR task unsupervised. The headset was delivered to them by courier. Participants received the links to the online follow-up assessments one, two, three and four months after they completed the baseline assessments.

### Data analysis

All data were analysed using IBM SPSS statistics version 27 and R studio version 4.1.3 Means and standard deviations were calculated for proportion of choosing each action tendency per condition (self-agency and other-agency) in the VR action tendency task and described in our previous study (Duan, Valmaggia, et al., 2022). A multiple regression analysis was conducted with all the pre-registered predictors in the primary analysis as independent variables and depressive symptoms as assessed by QIDS-SR-16 at four months’ follow-up as the outcome variable. The pre-registered predictors in the primary analysis include: 1) proportion of trials during which hiding was chosen as measured by the VR task; 2) proportion of trials during which distancing from oneself was chosen as measured by the VR task; 3) autonomy subscale score as measured by the Personal Style Inventory-II (PSI-II, Robins et al., 1994) 4) sociotropy subscale score as measured by the PSI-II; 5) Maudsley staging method (MSM) total score (Fekadu et al., 2018); 6) medication adherence during the four-month follow-up period as measured by a question (how regularly have you taken your antidepressants over the last month at the prescribed dose) where participants were asked to choose from: never, some of the time, more than half of the time, most of the time, almost every day, every day; 7) social support received as measured by the Social Support Scale (SSS, Krause & Borawski-Clark, 1995); 8) baseline depressive symptoms as measured by QIDS-SR-16; 9) baseline anxiety symptoms as measured by the GAD-7; 10) antidepressant changes during the four-month follow-up period where participants were classified into three categories from minor change to major change: no new antidepressant/stop current antidepressant/lower the dosage of current antidepressant, increase from effective dose to higher dose, increase from ineffective to effective dose/change to another antidepressant at effective dose.

Using a data-driven approach, to reduce the number of variables and avoid overfitting of our data-driven regression model, the proportions of all the maladaptive action tendencies and self-blaming rating biases as measured by the VR action tendencies task and the continuous clinical measures including subscales in the primary analysis were entered into a Principal Component Analysis (PCA) with varimax rotation. Factors extracted by the PCA were labelled and then used to predict depressive symptoms at four months’ follow-up along with all our ordinal clinical variables in the primary analysis: MSM total score, medication adherence and antidepressant changes. The number of factors to retain was first determined by inspection of the scree-plot using an elbow method [(Thorndike, 1953), see Supplementary Figure 1]. In addition, as suggested by Stevens (2009) to improve reproducibility, factors were only interpreted if the average of the four largest loadings was >.60 and only items with factor loadings above 0.51 were considered to load significantly on a factor.

In a further supporting analysis, to further understand the specific role of maladaptive action tendencies on prognosis, we selected “feeling like punishing oneself” as the action tendency which had loaded most strongly on one of the extracted PCA factors, and used a linear mixed model to examine its effect on the trajectory of depressive symptoms over the four monthly follow-up VQIDS-SR-5 scores. People with depression were sub-grouped based on whether they had or had not selected “feeling like punishing myself” at least once in the VR task at baseline, as binarizing the action tendency measure is what one would do in a clinical applied setting where people would not have factor scores. Action tendency group was then entered in the linear mixed model as a categorical independent variable, along with time and their interactions. Post-hoc analyses were conducted for significant interactions. Multiple comparison correction at a two-sided Bonferroni-corrected p=.05 was employed for all post-hoc univariate tests.

In addition, a doubly Cross-Validated Elastic-net regularised generalised linear model of the gaussian family (dCVnet, Lawrence et al., 2021) was performed to examine the cross-validated model performance when predicting the prognosis of depression as measured by depressive symptoms at final follow-up assessment. This method uses double cross-validation and regularisation to guard against overfitting and cross-validation, allowing a more accurate estimate of model performance which is more likely to generalise to independent future samples. Because of the built-in elastic-net regularisation of the dCVnet method, all VR-task predictors included in the PCA were included in the first dCVnet model without prior conversion to factors. In addition, all our ordinal clinical variables in the primary analysis that could not be included in the PCA (MSM total score, medication adherence and antidepressant changes) were also included in the first dCVnet model. A second dCVnet model was conducted including all the variables in the first dCVnet model and variables in the secondary analyses to compare model performance after including additional pre-registered clinical variables and so probe our pre-registered hypothesis 3. More information regarding dCVnet can be found in the Supplementary Methods.

## Results

### Pre-registered primary prospective prediction model

A multiple linear regression was conducted to predict depressive symptoms at four months’ follow-up based on all the pre-registered predictors in the primary analysis. A significant regression equation was found: F(11,77)=3.75, p<.001, with an R^2^ of .33. Higher MSM total scores and higher depressive symptoms at baseline predicted higher depressive symptoms at four months’ follow-up. No other predictors were found to be significant (see Table 1).

**Table 1.** Regression model predicting depressive symptoms at four months’ follow-up based on all pre-registered predictors Overall model: F(11,77)=3.75, p<.001, R^2^=.33; MSM=Maudsley staging method; QIDS-16=Quick Inventory of Depressive Symptomatology-16; GAD-7=Generalised Anxiety Disorder-7; PSI: Personal Style Inventory-II;* p<.05.

|  | Unstandardized Coefficients |  | Standardized Coefficients | t-value | p-value | **Table 1\|**<br>Regression model predicting depressive symptoms at four months' follow-up based on all pre-registered predictors |
| --- | --- | --- | --- | --- | --- | --- |
|  | B | Std. Error | Beta |  |  |  |
| Hide (VR task) | -2.91 | 6.68 | -0.05 | -0.44 | 0.664 |  |
| Distance from self (VR task) | 5.02 | 10.81 | 0.05 | 0.46 | 0.644 |  |
| Sociotropy subscale of the PSI | 0.01 | 0.04 | 0.04 | 0.32 | 0.746 |  |
| Autonomy subscale of the PSI | 0.06 | 0.04 | 0.16 | 1.47 | 0.147 |  |
| MSM total score | 1.00 | 0.46 | 0.27 | 2.20 | 0.031* |  |
| Medication adherence | -0.12 | 0.25 | -0.05 | -0.50 | 0.620 |  |
| Support received | 0.06 | 0.08 | 0.07 | 0.68 | 0.500 |  |
| QIDS-16 total score | 0.38 | 0.18 | 0.28 | 2.09 | 0.040* |  |
| GAD-7 total score | 0.02 | 0.12 | 0.02 | 0.15 | 0.882 |  |
| Antidepressant changes | -1.28 | 0.85 | -0.15 | -1.52 | 0.133 |  |

### Data-driven prospective prediction model based on all pre-registered predictors of interest

A principal component analysis with varimax rotation was conducted on all the continuous measures (including subscales) pre-registered for the primary analysis. The rotated component matrix is shown in Table 2. The elbow method of inspecting the scree-plot suggested a five-factor structure (See Supplementary Figure 1), which cumulatively explained 47.23% of the total variance. The factors were labelled according to their constitutive items as follows: Sociotropy/Perfectionism (factor 1, 4 items), Depressive and anxiety symptoms (factor 2, 3 items), Contact with friends (factor 3, 2 items), Response time (factor 4, 2 items) and Punishing oneself/self-blaming bias (factor 5, 4 items).

**Table 2.** Principal Component Analysis: Varimax rotated factor loading of variables Factor 1: Sociotropy/Perfectionism; Factor 2: Depressive/Anxiety symptoms; Factor 3: Contact with friends; Factor 4: Response time; Factor 5: Punishing oneself/self-blaming bias. PSI=Personal Style Inventory-II; MM-PHQ-9=Maudsley Modified-Patient Health Questionnaire-9; QIDS-16=Quick Inventory of Depressive Symptomatology-16; GAD-7=Generalised Anxiety Disorder-7; SSS=Social Support Scale; SA=self-agency; OA=other-agency; rt=response time. *=above critical value (0.51) for significance. Factors were only interpreted if the average of the four largest loadings is >0.6 Stevens, 2009.

| Variables | Factor 1 | Factor 2 | Factor 3 | Factor 4 | Factor 5 |
| --- | --- | --- | --- | --- | --- |
| PSI concern about others | 0.87* | 0.07 | 0.09 | 0.07 | 0.09 |
| PSI perfectionism | 0.62* | 0.09 | -0.12 | 0.20 | 0.26 |
| PSI pleasing others | 0.55* | 0.28 | -0.12 | 0.05 | -0.04 |
| PSI dependency | 0.54* | 0.06 | 0.33 | -0.23 | 0.02 |
| MM-PHQ-9 total baseline | 0.09 | 0.89* | 0.02 | 0.08 | 0.02 |
| QIDS-16 total baseline | -0.02 | 0.79* | -0.10 | 0.11 | 0.05 |
| GAD-7 total baseline | 0.18 | 0.71* | -0.33 | -0.04 | -0.04 |
| PSI defensive separation | -0.04 | 0.13 | -0.79* | 0.04 | 0.05 |
| SSS contact friend | 0.04 | -0.08 | 0.73* | 0.06 | 0.10 |
| rt SA actions | 0.04 | 0.06 | 0.05 | 0.91* | 0.01 |
| rt OA actions | 0.09 | 0.11 | -0.01 | 0.90* | 0.09 |
| OA punish self | 0.07 | 0.02 | 0.02 | 0.15 | 0.76* |
| SA punish self | 0.13 | 0.06 | 0.30 | -0.23 | 0.61* |
| OA self-blaming bias | 0.16 | -0.07 | -0.08 | 0.12 | 0.58* |
| SA self-blaming bias | 0.35 | 0.24 | 0.05 | -0.39 | 0.44 |
| SSS support provided | 0.09 | 0.09 | 0.00 | -0.07 | 0.12 |
| SSS support received | 0.05 | -0.22 | 0.35 | 0.05 | 0.02 |
| SSS negative interaction | 0.17 | 0.36 | -0.45 | -0.06 | 0.04 |
| PSI need for control | 0.22 | 0.09 | -0.36 | 0.20 | -0.05 |
| OA verbally attack friend | -0.14 | 0.01 | -0.06 | -0.27 | -0.30 |
| OA hide | 0.37 | -0.02 | 0.13 | -0.10 | -0.02 |
| SA apologise | 0.07 | -0.01 | 0.01 | -0.01 | -0.04 |
| SA hide | 0.34 | -0.01 | -0.20 | 0.14 | -0.23 |
| OA apologise | 0.04 | -0.12 | 0.21 | -0.01 | 0.12 |
| SSS satisfaction | -0.06 | -0.24 | 0.36 | 0.10 | 0.08 |
| OA distance from self | -0.14 | -0.02 | -0.03 | 0.08 | 0.32 |
| SSS contact family | 0.19 | -0.24 | 0.09 | 0.06 | -0.27 |
| SA distance from self | -0.39 | -0.01 | 0.30 | -0.05 | 0.01 |
| OA distance from friend | 0.00 | 0.04 | -0.10 | 0.03 | -0.07 |

A multiple linear regression was conducted to predict depressive symptoms at four months’ follow-up based on these five factors along with the pre-registered ordinal predictors: MSM total score, medication adherence and antidepressant changes during the past four months. A significant regression equation was found: F(5,83)=5.89, p<.001, R^2^=.35. A higher MSM total score, a higher depressive and anxiety symptom factor score and a higher maladaptive action tendencies/self-blaming biases factor score predicted higher depressive symptoms at four months’ follow-up (see Table 3).

**Table 3.** Regression model predicting depressive symptoms at four months’ follow-up based on the factor analysis and ordinal clinical variables Overall model: F(5,83)=5.89, p<.001, R^2^=.35; MSM=Maudsley staging method; **p<.01, * p<.05

|  | Unstandardized Coefficients |  | Standardized Coefficients | t-value | p-value | Table 3\| Regression model predicting depressive symptom |
| --- | --- | --- | --- | --- | --- | --- |
|  | B | Std. Error | Beta |  |  |  |
| MSM total score | 1.04 | 0.39 | 0.28 | 2.68 | 0.009** |  |
| Medication adherence | -0.16 | 0.25 | -0.06 | -0.66 | 0.509 |  |
| Antidepressant changes | -1.26 | 0.83 | -0.14 | -1.52 | 0.132 |  |
| Sociotropy/Perfectionism factor | 0.19 | 0.49 | 0.04 | 0.39 | 0.701 |  |
| Depressive/anxiety symptom factor | 1.59 | 0.56 | 0.30 | 2.83 | 0.006** |  |
| Contact with friends factor | -0.16 | 0.48 | -0.03 | -0.32 | 0.748 |  |
| Response time factor | 0.41 | 0.48 | 0.08 | 0.87 | 0.388 |  |
| Punishing oneself/self-blaming bias factor | 1.22 | 0.48 | 0.23 | 2.51 | 0.014* |  |

### Influence of the tendency to punish oneself on depressive symptoms over four months

Following the significant association between factor 5 (maladaptive action tendencies/ self-blaming biases) and depressive symptoms at four months’ follow-up, a linear mixed model further examined the relationship between maladaptive action tendencies, which were the two strongest items loading on factor 5 (punishing oneself in the self-agency and other-agency conditions) and depressive symptoms as assessed by monthly VQIDS-5 over the follow-up period. After controlling for baseline VQIDS-5 score, there was a significant interaction between time and punishing oneself in the other-agency condition: B=.43, t(241.21)=2.08, p=.038. No main effect of time, punishing oneself in both agencies, nor interaction between time and punishing oneself in the self-agency condition were found (see Figure 1). As shown by **Figure 1**, post-hoc analysis revealed that participants had significantly higher depressive symptoms in the group with the tendency to punish oneself in the other-agency condition compared to the group without such tendency at four months’ follow-up only: F(1,84)=6.45, p=.01, partial Eta Squared=0.07. Although a similar trend was observed at other time points, there were no significant differences on depressive symptoms between the groups at these time points: F(1,79)=1.26, p=.27 (Follow-up 1); F(1,76)=1.30, p=.13 (Follow-up 2); F(1,71)=2.54, p=.12 (Follow-up 3).

**Figure 1.**
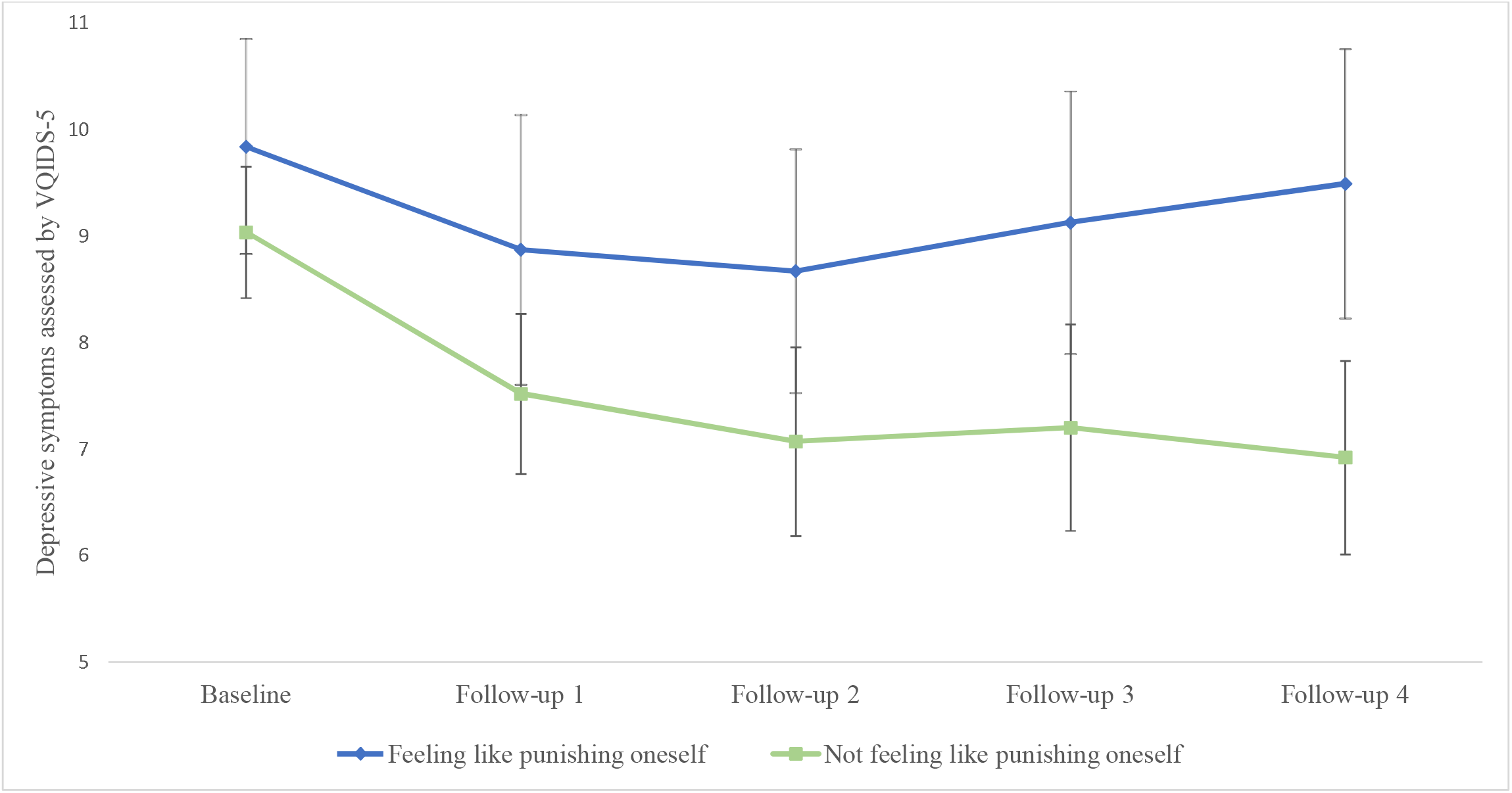
Trajectory of depressive symptoms in participants with and without agency-incongruent feeling like punishing themselves *Note*. This figure shows a significant interaction between time and punishing oneself in the other agency condition: B=.43, t(241.21)=2.08, p=.038. No main effect of time, punishing oneself in both agencies, nor interaction between time and punishing oneself in the self-agency condition were found. Post-hoc analysis revealed that participants had significantly higher depressive symptoms in the group with the tendency to punish oneself in the other-agency condition compared to the group without such tendency at 4 months’ follow-up only: F(1,84)=6.45, p=.01, (Partial) Eta Squared=0.07.

### Prediction of prognosis based on dCVnet models

Supporting our results of the simple linear regression model, four predictors were retained in the first dCVnet model (model with clinically established variables in our primary analysis): QIDS-SR-16, MM-PHQ9, MSM score and punishing oneself in the other-agency condition. Model performance of the two dCVnet models can be found in the Supplementary Results and shows that a cross-validated 19% of the variance in depressive symptoms after four months can be predicted when using all pre-registered primary and secondary variables.

## Discussion

The present study used a validated novel VR task to examine the role of maladaptive self-blame-related action tendencies in prognosis of current depression when treated as usual in primary care. Our pre-registered general hypothesis on the prognostic relevance of maladaptive action tendences was confirmed: punishing oneself for other people’s wrongdoing was associated with a poor prognosis of depression after four months. However, our findings did not support the more specific predictions that feeling like hiding and creating a distance from oneself would be prognostically relevant. We also examined the influence of clinically established risk factors. As expected, higher treatment resistance and more severe symptoms at baseline were associated with poor prognosis. Notably, none of the other clinical variables had significant prognostic value in this sample. Our statistical/machine learning model also identified the same three risk factors found using standard statistics and further confirmed that despite their significant contribution, useful individual level prediction of prognosis was not achieved using these measures alone.

Our previous cross-sectional study, in this sample of participants with current depression, found that punishing oneself for other people’s wrongdoing was the most clinically relevant action tendency in that it not only distinguished people with and without depression, but was also related to higher rates of self-harm (Duan, Valmaggia, et al., 2022). The present prospective study, despite not being informed by the above cross-sectional results at the pre-registration and analysis stage, further highlights the pathophysiological importance of an agency-incongruent feeling like punishing oneself by showing its selective predictive value for prognosis. This result was contrary to our more specific pre-registered hypotheses on feeling like hiding and creating a distance from oneself, which had been shown to be relevant to depression vulnerability in our previous studies in remitted depression (Duan, Valmaggia, et al., 2022; Lawrence et al., 2021). This may be due to the fact, that our previous studies were based on less immersive text-based measures, but may also indicate differential importance of specific action tendencies depending on symptomatic state and clinical subtype. This is because our previous studies selected fully remitting forms of depression rather than the more chronic difficult-to-treat population included in the current study.

In “Mourning and Melancholia”, Freud emphasized the role of internalised aggressive impulses such as self-attack/punishment in depression and differentiated depression from mourning on this basis (Freud, 1917). This tendency was thought to be a core feature of depression in Freudian models (Freud, 1917), but our results are more compatible with a model of depression in which a tendency to self-punish characterises a subgroup of depression. Furthermore, we share Beck’s criticism of the Freudian model in that the observation of self-punishing tendencies does not necessarily reflect a need to punish oneself (Beck, 1985). On a more cautionary note, we used an explicit measure of self-punishing tendencies, whereas an implicit measure would be more suitable to probe the Freudian model. Compared with self-punishment, hiding and creating a distance from oneself as precursors of social actions could give rise to maladaptive coping behaviours such as social avoidance (Mu & Berenbaum, 2019), which would explain their more prominent role in remitted depression at risk of recurrence. Interestingly, only punishing oneself in the other-agency, but not self-agency condition predicted prognosis of depression. This confirms theories and empirical findings that only overgeneralised forms of self-blame (e.g. punishing oneself for other people’s wrongdoing) contributes to the onset and maintenance of depression (Abramson et al., 1978; Janoff-Bulman, 1979) and confirms our finding of agency-incongruent self-blaming emotions in difficult-to-treat depression (Jaeckle et al., 2021).

We also found that depression severity and treatment resistance at baseline predicted the prognosis of depression after four months, confirming the importance of treatment history (Taylor et al., 2021; Webb et al., 2020). On the other hand, despite the effort of clinicians to increase medication adherence and social support among patients with depression, a higher score in these domains did not contribute to a better outcome. In addition, participants’ antidepressant changes during the follow-up period did not add value to the prediction, although this could be partially explained by the fact that very few participants changed their antidepressant medication overall, a reflection of the large treatment gaps in primary care for depression (Strawbridge et al., 2022).

Our study was limited by not including a diagnostic interview, so we were unable to establish a formal diagnosis for our participants. However, this makes our findings more generalisable to primary care patients where a formal diagnostic interview is not conducted. In addition, a highly conservative PHQ-9 score cut-off (≥ 15) was used in the study, with a specificity of .96 for MDD (Manea et al., 2012). We also used validated tools to exclude participants on the bipolar spectrum and those with alcohol or drug abuse.

Taken together, our study confirmed our general hypothesis that maladaptive self-blame-related action tendencies play a significant role in prognosis of current depression and were the only factor of prognostic relevance apart from well-known baseline levels of depressive symptoms and treatment-resistance. Our more specific pre-registered predictions about specific action tendencies were not confirmed, instead we found an overgeneralised feeling like punishing oneself and blaming oneself for other people’s wrongdoing as of distinctive prognostic importance for depression. We further showed that a simple binary categorisation of feeling like punishing oneself on our VR-task can be used to identify a subgroup of patients with poorer prognosis which could be used for personalising treatments and as a target for novel interventions.

## Data Availability

All data produced in the present study are available upon reasonable request to the authors

## Author Contributions

SD, RZ and VL developed the study concept and design. JM made significant contributions to the study concept. Data was collected by SD and DF. Testing and analysis were performed by SD under the supervision of RZ and AL. SD drafted the manuscript, and AL, LV, DF, JM and RZ provided critical revisions. RZ edited the final version. All authors approved the final version of the manuscript for submission.

## Acknowledgement

We greatly thank Jerome Di Pietro for the software development of the VR assessment in this study and are grateful to Phillippa Harrison for her advice on study instruments.

## Funding

This study was funded by a Scients Institute Catalyst Award to SD, who is also partly funded by a Henry Lester Trust Award and the Great Britain-China Educational Trust Chinese Student Award. DF was funded by the Medical Research Council Doctoral Training Partnership (ref: 2064430). AW, RZ, LV and the VR Lab were partly funded by the National Institute for Health and Care Research (NIHR) Biomedical Research Centre at South London and Maudsley NHS Foundation Trust and King’s College London. The views expressed are those of the authors and not necessarily those of the NHS, the NIHR or the Department of Health and Social Care.

## Conflicts of interest

RZ is a private psychiatrist service provider at The London Depression Institute and co-investigator on a Livanova-funded observational study of Vagus Nerve Stimulation for Depression. RZ has received honoraria for talks at medical symposia sponsored by Lundbeck as well as Janssen. He has collaborated with EMOTRA, EMIS PLC and Depsee Ltd. He is affiliated with the D’Or Institute of Research and Education, Rio de Janeiro and advises the Scients Institute, USA. The other authors have no conflicts of interests to declare.

## Notes

### Author Declarations

This study was approved by the King's College London PNM Research Ethics Subcommittee (Project Reference:HR-19/20-17589)

