## Supplementary materials for "Virtual reality-assessment of social interactions and prognosis in depression"

**Supplementary Methods**

***Prediction of prognosis of depression using dCVnet***

The R package “dCVnet” [https://www.github.com/ andrewlawrence/dCVnet](https://www.github.com/%20andrewlawrence/dCVnet) was used as planned in our pre-registered analysis plan (Lawrence et al., 2021). dCVnet uses a nested cross-validation scheme to simultaneously select hyperparameters which cross-validate well (the inner loop) and obtain uninflated cross-validated performance estimates (the outer loop). For these models stable hyper-parameter selection was obtained with 30 repetitions of 10-fold cross-validation in the inner loop. Stable cross-validated performance estimates were obtained with 100 repetitions of 10-fold cross-validation in the outer loop. Both alpha (type of penalty) and lambda (amount of penalty) were tuned. Six logarithmically spaced values of alpha were considered between 0.01 (mostly Ridge) and 1.0 (a LASSO model). For each alpha, 100 lambda values were determined automatically, logarithmically spaced between the lambda giving a fully penalised model and 0.0001. The hyper-parameters (Alpha and Lambda) were selected based on the minimum mean square error. More information regarding the methods can be found in Lawrence et al. (2021).

***Inclusion and Exclusion of participants***

Full details of the inclusion and exclusion reasons of participants can be found in the Supplementary Materials of Duan et al. (2022).

**Supplementary Table 1|** Demographic characteristics of participants at baseline

| Age | 32.05 ±12.32 (18-66) |
| --- | --- |
| Years of Education | 16.40 ±2.97 (4-22) |
| Sex [female] | 89 (85.6%) |
| Native language |  |
| *English* | 87 (86.1%) |
| *Other* | 14 (13.9%) |
| Employment status |  |
| *In full-time employment* | 26 (25.7%) |
| *In part-time employment* | 11 (10.9%) |
| *Retired* | 2 (2%) |
| *Student* | 37 (36.6%) |
| *Unemployed* | 13 (12.9%) |
| *Other* | 12 (11.9%) |
| Ethnicity |  |
| *Asian or Asian British: Bangladeshi* | 4 (4%) |
| *Asian or Asian British: Chinese* | 1 (1%) |
| *Asian or Asian British: Indian* | 3 (3%) |
| *Asian or Asian British: Pakistani* | 2 (2%) |
| *Asian or Asian British: Other* | 1 (1%) |
| *Black or Black British: African* | 0 |
| *Black or Black British: Caribbean* | 1 (1%) |
| *Black or Black British: Other* | 3 (3%) |
| *Mixed: White & Black Caribbean* | 2 (2%) |
| *Mixed or Multiple: Other* | 3 (3%) |
| *White: British* | 64 (63.4%) |
| *White: Irish* | 2 (2%) |
| *White: Other* | 9 (8.9%) |
| *Other* | 6 (5.9%) |

Values are Mean ± Standard Deviation (range) for continuous variables and Count (Percentage%) for categorical variable. Sample size was n=101.

| PHQ-9 total score  at pre-screening | 18.86±3.07(15-27) |
| --- | --- |
| MM-PHQ-9 total score  at baseline | 18.08±4.75(5-27) |
| QIDS-SR-16 total score  at baseline | 16.90±4.07(5-26) |
| GAD-7 total score at baseline | 12.14±5.44 (0-21) |
| Number of failed treatments |  |
| *1-2* | 61 (60.4%) |
| *3-4* | 34 (33.7%) |
| *5-6* | 5 (5%) |
| *7-10* | 1 (1%) |
| Duration of current depressive episode |  |
| *≤12 months* | 76 (75.2%) |
| *13-24 months* | 10 (9.9%) |
| *>24 months* | 15 (14.9%) |
| Age at first onset | 16.41±6.48 (4-55) |
| Maudsley staging method total score | 6.47±1.40 (4-11) |
| Maudsley staging method severity |  |
| *Mild* | 60 (57.7%) |
| *Moderate* | 40 (38.5%) |
| *Severe* | 1 (1%) |
| Self-reported co-morbid psychiatric conditions |  |
| *PTSD* | 10 (10.0%) |
| *Anxiety disorders* | 7 (6.9%) |
| *Eating disorders* | 7 (6.9%) |
| *Personality disorders* | 5 (4.9%) |
| *OCD* | 5 (4.9%) |
| *ASD* | 3 (3.0%) |
| *ADHD* | 1 (1.0%) |
| *Other* | 4 (4.0%) |
| Current medication |  |
| *Single SSRI* | 58 (55.8%) |
| *Single SNRI* | 13 (12.5%) |
| *Other* | 14 (13.4%) |
| *None* | 19 (18.3%) |

**Supplementary Table 2|** Clinical characteristics of participants at baseline

QIDS-SR-16: The Quick Inventory of Depressive Symptomatology-Self-Report-16; MM-PHQ9: Maudsley Modified Patient Health Questionnaire-9; GAD-7: General Anxiety Disorder-7. SSRI: Selective serotonin reuptake inhibitors; SNRI: Serotonin and norepinephrine reuptake inhibitors. PTSD: posttraumatic stress disorder; OCD: obsessive-compulsive disorder; ASD: autism spectrum disorder; ADHD: attention deficit hyperactivity disorder; Values are Mean ± Standard Deviation (range) for continuous variables and Count (Percentage%) for categorical variable. Sample size was n=101.

**Supplementary Table 3|** Coefficients in the final model of dCVnet for all variables in the primary analysis

| Predictor | Final Model | Outer Median | min | max |
| --- | --- | --- | --- | --- |
| PSI concern about others | n.s. | 0.000 | -0.181 | 0.051 |
| PSI dependency | n.s | 0.000 | -0.006 | 0.151 |
| PSI pleasing others | n.s | 0.002 | 0.000 | 0.274 |
| PSI perfectionism | n.s | 0.004 | 0.000 | 0.289 |
| PSI need for control | n.s | 0.048 | 0.000 | 0.471 |
| PSI defensive separation | n.s | 0.043 | 0.000 | 0.607 |
| SA punish self | n.s | 0.000 | 0.000 | 0.220 |
| SA apologize | n.s | 0.000 | -0.013 | 0.139 |
| SA hide | n.s | -0.030 | -0.560 | 0.000 |
| SA distance from self | n.s | -0.003 | -0.302 | 0.000 |
| OA verbally attack friend | n.s | -0.013 | -0.427 | 0.000 |
| OA punish self | 0.163 | 0.172 | 0.000 | 0.535 |
| OA apologize | n.s | 0.000 | -0.048 | 0.069 |
| OA hide | n.s | 0.000 | -0.064 | 0.069 |
| OA distance from self | n.s | 0.029 | 0.000 | 0.470 |
| OA distance from friend | n.s | 0.000 | -0.029 | 0.156 |
| QIDS total baseline | 0.811 | 0.841 | 0.104 | 1.855 |
| MM-PHQ-9 total baseline | 0.535 | 0.538 | 0.000 | 1.388 |
| GAD-7 total baseline | n.s | 0.000 | 0.000 | 0.231 |
| SSS contact family | n.s | 0.000 | -0.315 | 0.000 |
| SSS contact friend | n.s | 0.017 | 0.000 | 0.549 |
| SSS support received | n.s | 0.000 | -0.016 | 0.223 |
| SSS support provided | n.s | 0.004 | 0.000 | 0.317 |
| SSS negative interaction | n.s | 0.000 | -0.131 | 0.146 |
| SSS satisfaction | n.s | 0.000 | -0.103 | 0.071 |
| OA self-blaming bias | n.s | 0.028 | 0.000 | 0.404 |
| SA self-blaming bias | n.s | 0.015 | 0.000 | 0.439 |
| rt SA actions | n.s | 0.000 | -0.091 | 0.211 |
| rt OA actions | n.s | 0.014 | 0.000 | 0.397 |
| MSM total score | 0.536 | 0.539 | 0.000 | 0.991 |
| Medication adherence | n.s | 0.000 | -0.318 | 0.000 |
| Antidepressant changes | n.s | -0.054 | -0.702 | 0.000 |

PSI=Personal Style Inventory-II; MM-PHQ-9=Maudsley Modified-Patient Health Questionnaire-9; QIDS-16=Quick Inventory of Depressive Symptomatology-16; GAD-7=Generalised Anxiety Disorder-7; SSS=Social Support Scale; SA=self-agency; OA=other-agency; rt=response time; MSM=Maudsley Staging Model. Final Model: coefficients in the final/production model; Outer Median: the median of the coefficients over the outer loop of the cross-validation. n.s.= not selected

**Supplementary Table 4|** Outer-loop performance of dCVnet model including all the variables in the primary analysis

| Measure | mean | SD | min | max |
| --- | --- | --- | --- | --- |
| RMSE | 4.830 | 0.069 | 4.673 | 5.023 |
| MAE | 3.945 | 0.061 | 3.810 | 4.102 |
| r | 0.377 | 0.045 | 0.265 | 0.471 |
| r^2^ | 0.143 | 0.030 | 0.070 | 0.222 |
| cal Intercept | -2.512 | 2.232 | -8.389 | 2.937 |
| cal Slope | 1.169 | 0.152 | 0.792 | 1.570 |
| Brier | 23.329 | 0.666 | 21.835 | 25.230 |
| SDScaledRMSE | 0.923 | 0.013 | 0.893 | 0.960 |
| SDScaledMAE | 0.754 | 0.012 | 0.728 | 0.784 |

RMSE=root mean square deviation; MAE=mean absolute error; cal=calibration; SD=standard deviation; SDScaledRMSE = RMSE divided by the outcome standard deviation; SDScaledMAE = MAE divided by the outcome standard deviation.

**Supplementary Table 5|** Outer-loop performance of dCVnet model including all the variables in the primary and secondary analysis

| Measure | mean | SD | min | max |
| --- | --- | --- | --- | --- |
| RMSE | 4.706 | 0.078 | 4.545 | 4.882 |
| MAE | 3.783 | 0.0753 | 3.603 | 3.958 |
| r | 0.43 | 0.0342 | 0.363 | 0.487 |
| r^2^ | 0.186 | 0.0248 | 0.132 | 0.237 |
| cal Intercept | 1.21 | 1.2657 | -1.887 | 4.074 |
| cal Slope | 0.914 | 0.0866 | 0.722 | 1.12 |
| Brier | 22.156 | 0.7343 | 20.659 | 23.836 |
| SDScaledRMSE | 0.899 | 0.0149 | 0.869 | 0.933 |
| SDScaledMAE | 0.723 | 0.0144 | 0.689 | 0.756 |

RMSE=root mean square deviation; MAE=mean absolute error; cal=calibration; SD=standard deviation; SDScaledRMSE = RMSE divided by the outcome standard deviation; SDScaledMAE = MAE divided by the outcome standard deviation.

**Supplementary Figure 1|** Scree Plot of components in the principal component analysis including all the variable in the primary analysis except the ordinal variables

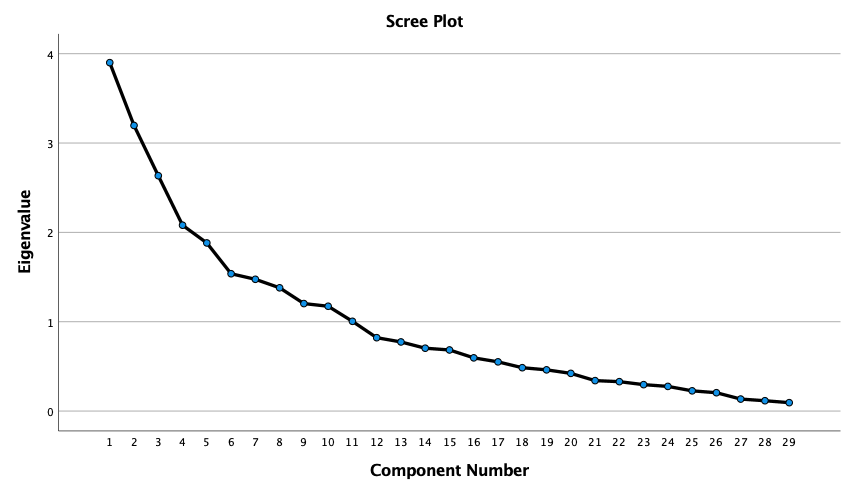
